# Genomic and molecular associations with preoperative immune checkpoint inhibition in patients with stage III clear cell renal cell carcinoma

**DOI:** 10.1101/2025.07.31.25332518

**Authors:** Wesley H. Chou, Lucy Lawrence, Emma Neham, Shreeram Akilesh, Amy E. Moran, Christopher L. Corless, Lisa Langmesser, Beyza Cengiz, Kazumi Eckenstein, Jen-Jane Liu, Sudhir Isharwal, Christopher L. Amling, Marshall C. Strother, Nicholas H Chakiryan, George V. Thomas

**Author notes:** The last two authors are co-senior authors. **Author emails:** LL EN SA AEM CLC LL BC KE JL SI CLA MCS GVT NHC. **Corresponding author:** Wesley H. Chou, MD, Oregon Health & Science University, 3181 SW Sam Jackson Park Road, Portland, Oregon 97202, United States of America.

## Abstract

**Purpose:** Patients with stage III clear cell renal cell carcinoma are at high risk for recurrence after nephrectomy. To mitigate overtreatment, there is a pressing clinical need to determine which patients benefit most from perioperative immune checkpoint inhibition. We performed a multimodal digital spatial analysis of gene and protein expression in stage III primary renal cell carcinomas, a subset of which had preoperative immune checkpoint inhibition exposure.

**Materials and Methods:** Surgically resected tumors from stage III clear cell renal cell carcinoma patients were analyzed using the Nanostring GeoMx Digital Spatial Profiler. Differential expression analysis was performed and validated using NCT02210117 trial data to identify genes associated with immune checkpoint blockade and clinical response. A gene score was then generated to predict overall survival in patients from The Cancer Genome Atlas.

**Results:** Among 19 patients, RNA expression significantly differed based on preoperative immune checkpoint blockade and recurrence – CD8+ effector and central-memory T-cell signatures were less prevalent in the treatment-naïve with recurrence group. Three out of four patients with preoperative immune checkpoint inhibition had recurrence. External validation yielded a 4-gene set (*GZMK, GZMA, ITGAL*, and *IL7R*); higher gene expression levels predicted better overall survival in The Cancer Genome Atlas cohort (p=0.005).

**Discussion:** Preoperative immune checkpoint blockade favorably altered the tumor microenvironment to resemble that of treatment-naïve patients without recurrence. However, this did not translate to better clinical outcomes. On external validation, the genes *GZMK, GZMA, ITGAL*, and *IL7R* were modifiable with immune checkpoint inhibition and associated with improved survival. Further investigation to assess if patients with low baseline expression of these genes may particularly benefit from perioperative immune checkpoint blockade is warranted.

## Introduction

Patients with stage III clear cell renal cell carcinoma (ccRCC) are at a high risk of disease recurrence after surgical resection – historically between 30-50%.^1^ With this in mind, there has been substantial effort to develop new treatment strategies incorporating immune checkpoint inhibitors (ICIs) to minimize recurrence risk in these patients.^2^ From this effort, the sole positive study has been KEYNOTE-564, a phase 3 double-blind randomized controlled trial published in 2021, which found that adjuvant pembrolizumab significantly improved disease-free survival versus placebo in patients with ccRCC at a high risk for recurrence (hazard ratio [HR] 0.72, 95% confidence interval [CI] 0.59-0.87).^3^ Based on these results, current National Comprehensive Cancer Network (NCCN) guidelines for kidney cancer recommend adjuvant pembrolizumab versus surveillance for surgically resected stage III ccRCC.^4^ In contrast, more recent results from the PROSPER trial failed to show clinical benefit from pre- and post-operative nivolumab in a similar patient cohort (HR 0.94, 95% CI 0.74-1.21).^5^

Clinically, there remains a pressing need to determine which stage III ccRCC patients will benefit from perioperative ICI therapy -- avoiding treatment for patients who can be cured with surgery alone and reserving treatment for patients who stand to derive the most benefit. Additionally, there is little consensus on what effect ICIs have on the tumor immune microenvironment of stage III ccRCC primary cancers and what these changes portend for subsequent clinical outcomes. To work toward these ends, we performed a multimodal digital spatial analysis of gene and protein expression in primary tumors from patients with stage III ccRCC, a subset of whom had preoperative ICI exposure – with additional refinement, validation, and computational analysis using several independent cohorts.

## Materials and Methods

### Tumor samples

Patients initially diagnosed with stage III ccRCC without sarcomatoid or rhabdoid histology with surgical resection of primary kidney tumors between 2016-2021 were identified in the prospectively collected OHSU Knight Cancer Center Bio-library. All patients were prospectively consented for data and specimen collection and subsequent analysis. Approval was obtained from the local institutional review board (OHSU-IRB #4918).

Patients either had ≥48 months of documented recurrence-free postoperative follow-up or biopsy-proven clinical recurrence within that timeframe. Three main groups were used for analysis: patients treated with preoperative PD-1 inhibiting ICI as monotherapy or in combination with a VEGF tyrosine kinase inhibitor as clinically indicated for tumor downstaging (preoperative ICI); patients without preoperative ICI with clinical recurrence (treatment-naïve with recurrence); patients without preoperative ICI without clinical recurrence (treatment-naïve without recurrence). No patients received adjuvant therapy.

### Gene and protein expression analysis

Tumor microarrays (TMAs) were constructed from multiple 1 mm diameter cores from each formalin-fixed and paraffin-embedded (FFPE) surgically resected primary tumor sample. Within each TMA core, 400-660 µm diameter circular regions of interest (ROI) were analyzed using Nanostring GeoMx Digital Spatial Profiler (DSP) protein analysis (an 87-protein oncology-specific panel), and Nanostring GeoMx DSP Human Whole Transcriptome Atlas RNA analysis (>18,000 genes).^6,7^ The selected ROIs for analysis were concentrated areas of tumor cells without significant intra-tumoral necrosis, major vasculature, or processing-related artifact or fracturing. Specimens and regions of interest were processed and analyzed using manufacturer pre-specified protocols.^8^

Briefly, reagents consisted of a suspension of DSP oligonucleotide barcodes attached via a photocleavable linker to either target-specific antibodies for protein analysis, or target-specific complementary nucleotide sequences for RNA analysis. The TMA tissue was then stained with imaging probes and assay probes, using either the protein or RNA reagent. Barcodes from the selected regions of interest were released via ultraviolet light exposure and quantified downstream via the nCounter analysis system.^9^ For protein analysis, raw counts were normalized using the geometric mean of housekeeping proteins and immunoglobulin controls, per manufacturer recommended protocols. For RNA analysis, proprietary positive and negative controls are introduced to ensure quality control and sample integrity and Q3 normalization is performed, per manufacturer recommended protocols, resulting in similar gene expression ranges for all samples.

The normalized counts of each region were analyzed individually for heterogeneity and averaged on a per-patient level for each region analyzed for each individual patient. The DSP protein data was filtered for 15 proteins relevant to immune cell phenotype and function. After quality control steps filtering out genes with very low expression or abnormally high variance, the DSP RNA data included 16,173 genes. RNA data was normalized using *voom* to estimate the mean-variance relationship and generate precision weights for subsequent differential expression analysis with *limma*.^10,11^ Enrichment analysis using the *Reactome* 2024 gene set was applied to clinically relevant sets of differentially expressed genes.^12^ *xCell* immune cell deconvolution was applied to the normalized RNA data to estimate the relative immune cell content between samples in our study.^13^

Heatmaps were generated for the protein, RNA, and xCell data using *pheatmap*, stratifying patients by the following groups: preoperative ICI, treatment-naive without recurrence, and treatment-naive with recurrence.^14^ Jittered plots were generated for protein, RNA, and xCell results demonstrating false discovery adjusted (FDR)-adjusted non-parametric statistical significance between treatment groups. Spearman’s correlations were determined for matched pairs of genes and their protein products.

### External validation and generation of gene score for prognostication

We refined and validated our differential expression analysis using RNA-seq data from the NCT02210117 randomized clinical trial (Goswami et al. 2025) that randomly assigned patients to one of three ICI-containing treatment regimens and obtained either surgically resected or biopsy tissue post-ICI exposure.^15^ With the goal of identifying genes that were both 1) associated with favorable clinical outcomes and 2) modifiable with ICI, differentially expressed genes associated with non-recurrence and ICI-exposure from our initial experiment were utilized as an initial set. These genes were matched with genes available in the RNA analysis of matched pre-treatment and ICI-exposed tumor tissue from the NCT02210117 trial, which reported 737 genes from the Nanostring Pan-cancer Immune Profiling assay. Patients were included if they had matched pre- and post-ICI tissue for analysis. The resultant matched genes were then filtered using clinical outcomes from the trial, selecting genes that demonstrated statistically significant increased expression after ICI-exposure, then further selecting genes whose increased post-ICI expression was associated with objective response to treatment, per RECIST criteria.^16^

To determine which cell types are likely to express these genes, the final gene set after validation/refinement with NCT02210117 data. (*GZMA, GZMK, IL7R, ITGAL*) was assessed using a meta-analysis of scRNA-seq data derived from 14 experiments on tumor tissue from patients with ccRCC, facilitated by *UncoVer*.^17^ Briefly, uniform manifold approximation and projection (UMAP) was applied to normalized and integrated scRNA-seq data from the included studies, then broadly-generalizable clusters of immune, stromal, and tumoral cells were identified using canonical gene markers. Expression levels of each gene in our final set were plotted to determine which cell types are likely to express these genes.

The final gene set after validation/refinement with NCT02210117 data was used to create a gene score algorithm (*GZMA, GZMK, IL7R, ITGAL*) using the geometric mean of each included gene. This score was applied to treatment-naïve clinical stage III patients in The Cancer Genome Atlas clear cell RCC (TCGA KIRC) cohort (n=123) to determine associations with overall survival. The TCGA was accessed using the *TCGAbiolinks* package. Patients were stratified by the median value of the gene score into high and low scores, then Kaplan-Meier survival curves were generated.

### Statistical analysis

All downstream computational and statistical analysis was conducted using R version 4.4.2 (R Foundation for Statistical Computing). Statistical significance was determined using non-parametric FDR-adjusted p < 0.05. Kruskal-Wallis rank sum testing was utilized to test statistical differences between the three clinical categories. When appropriate, Wilcoxon signed rank or rank sum testing was used to test statistical differences when two groups were present. Survival analysis was conducted using the *survival* and *survminer* packages.

## Results

Nineteen patients with stage III ccRCC were identified and included in our analysis; basic characteristics of the cohort are described in **Supplemental Table 1**. Four patients received preoperative PD-1-inhibiting ICI therapy, of which three had a clinical recurrence during the follow-up period. Among the 15 patients who did not receive preoperative ICI, eight experienced a recurrence and seven were disease-free with ≥48 months of documented follow up.

Among the 19 specimens, 41 cores were obtained to create a TMA (2-3 cores per specimen). Among the 41 cores, 41 RNA ROIs (1 per core), and 54 protein ROIs (1-3 per core), were selected for GeoMx DSP analysis. A diagram of the experimental workflow is depicted in **Fig. 1**. Examples of how digital spatial profiling was leveraged to accurately select concentrated tumor ROIs are shown in **Fig. 2A**.

**Figure 1.**
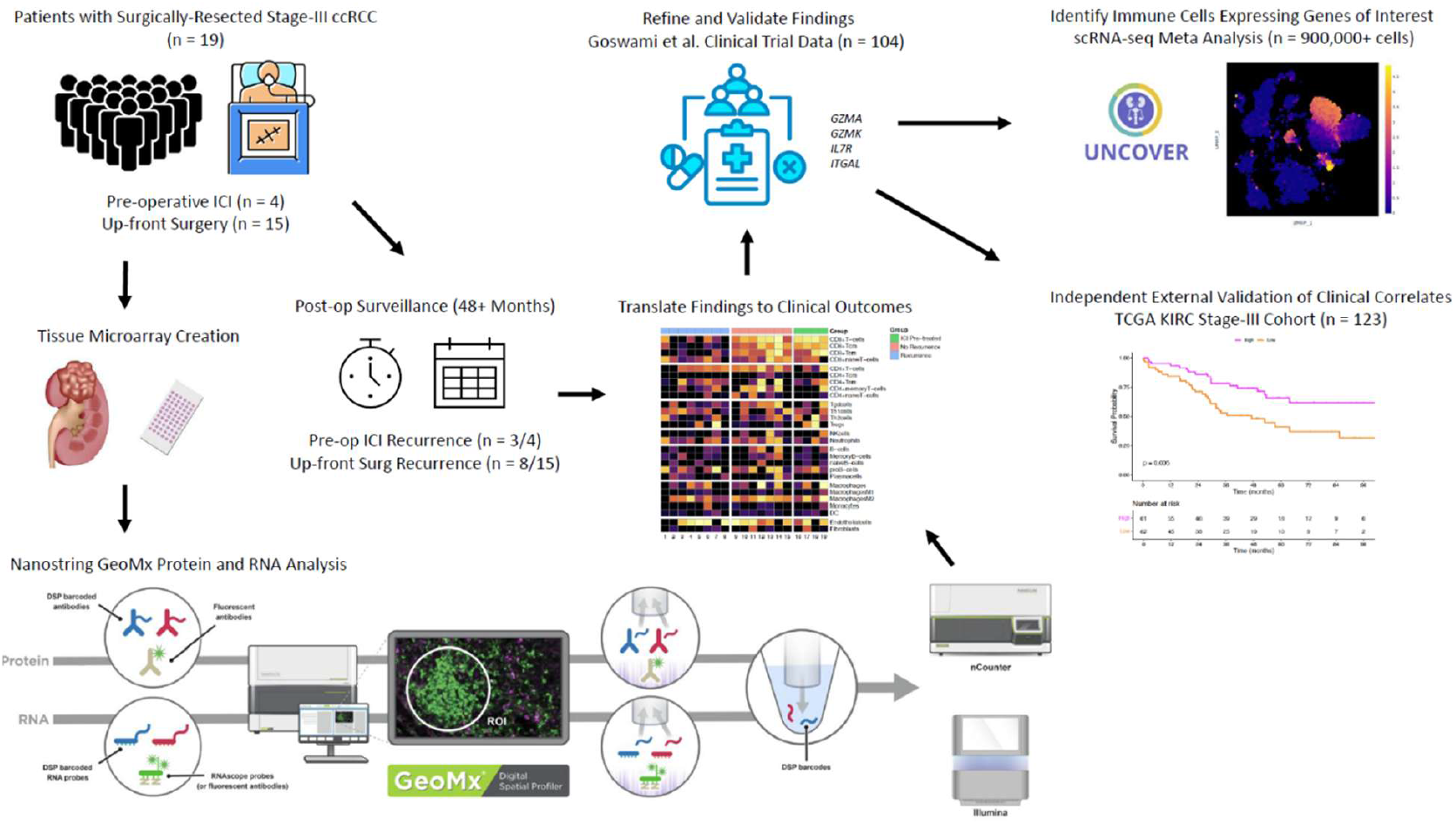
Schematic of study design and workflow.

**Figure 2.**
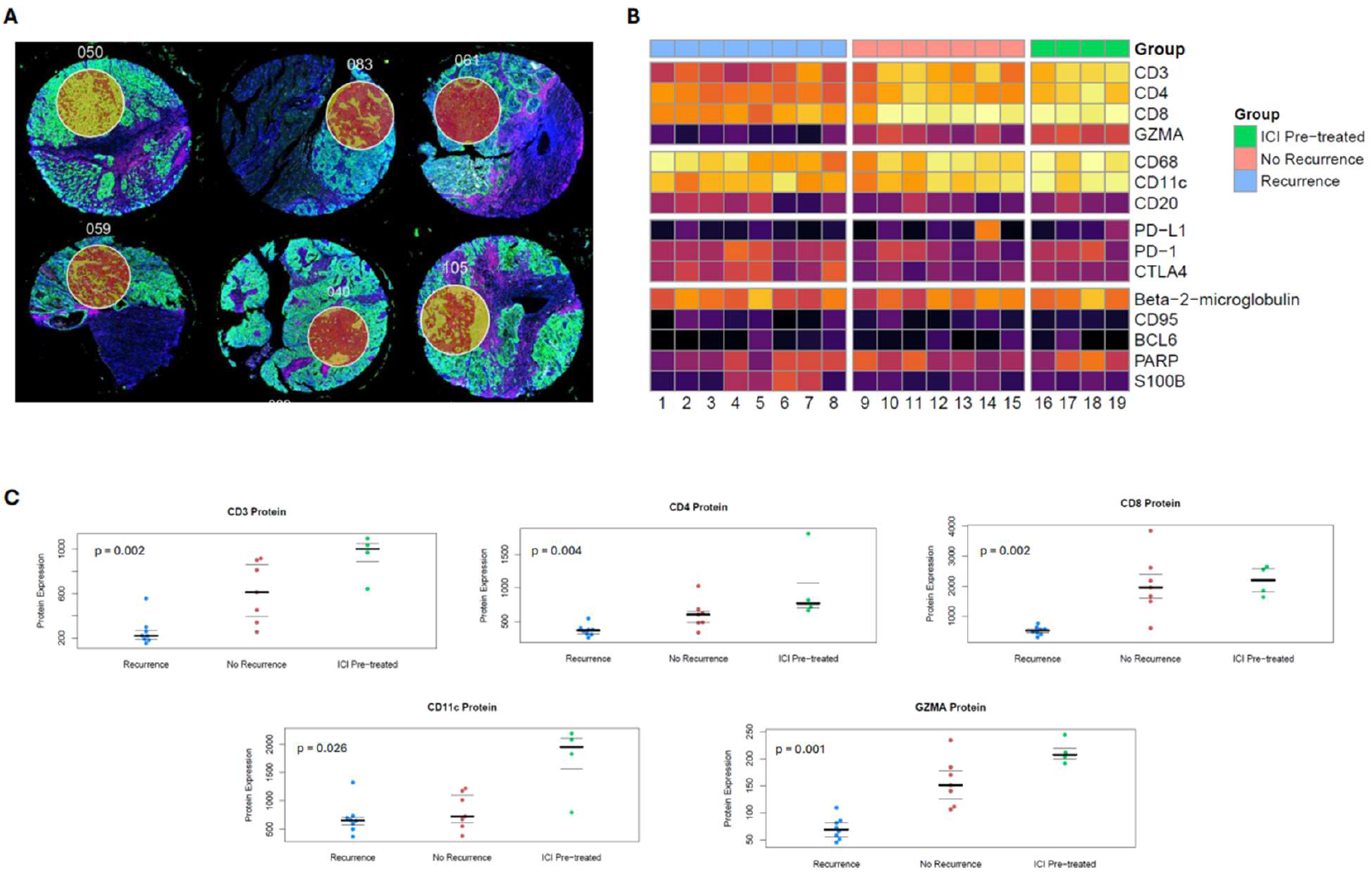
(A) Examples of ROI selection using Nanostring GeoMx Digital Spatial Profiler post-staining, pre-cleaving, which enables precise sampling of concentrated tumoral regions, avoiding areas of necrosis, major vasculature, tissue artifact, or normal adjacent kidney. (B) Heatmap for expression of 15 proteins relevant to immune cell function for patients who were treatment-naïve with recurrence (blue), treatment-naïve without recurrence (pink), or with preoperative ICI exposure (green). (C) Jittered plots for protein expression where protein expression significantly differed between the three groups, which included CD3, CD4, CD8, CD11c, and GZMA. The bold line represents the median and thin lines represent the interquartile range.

### Protein marker analysis

Protein expression of CD3, CD4, CD8, GZMA, and CD11c was significantly different between the three clinical groups: preoperative ICI exposure, treatment-naïve without recurrence, and treatment-naïve with recurrence (**Fig. 2B**). Higher expression of these proteins was seen in tumor samples from patients with preoperative ICI or treatment-naïve non-recurrence when compared to treatment-naïve patients with recurrence (**Fig. 2C)**. Notably, the immune checkpoints PD-1, PD-L1, and CTLA-4 did not demonstrate statistically significant differences between groups.

### Gene expression analysis

RNA expression data was filtered for the most variably expressed genes between groups (**Fig. 3A)**. Normalized log2 RNA expression was significantly different between the 3 groups for *CD3D, CD8A, GZMA, GZMK, ITGAL, IL7R, MIDN, TMEM9*, and *MYOCOS* (**Fig. 3B)**, as well as several other genes reported in **Supplemental Table 2** that were omitted from Fig. 3 for brevity.

**Figure 3.**
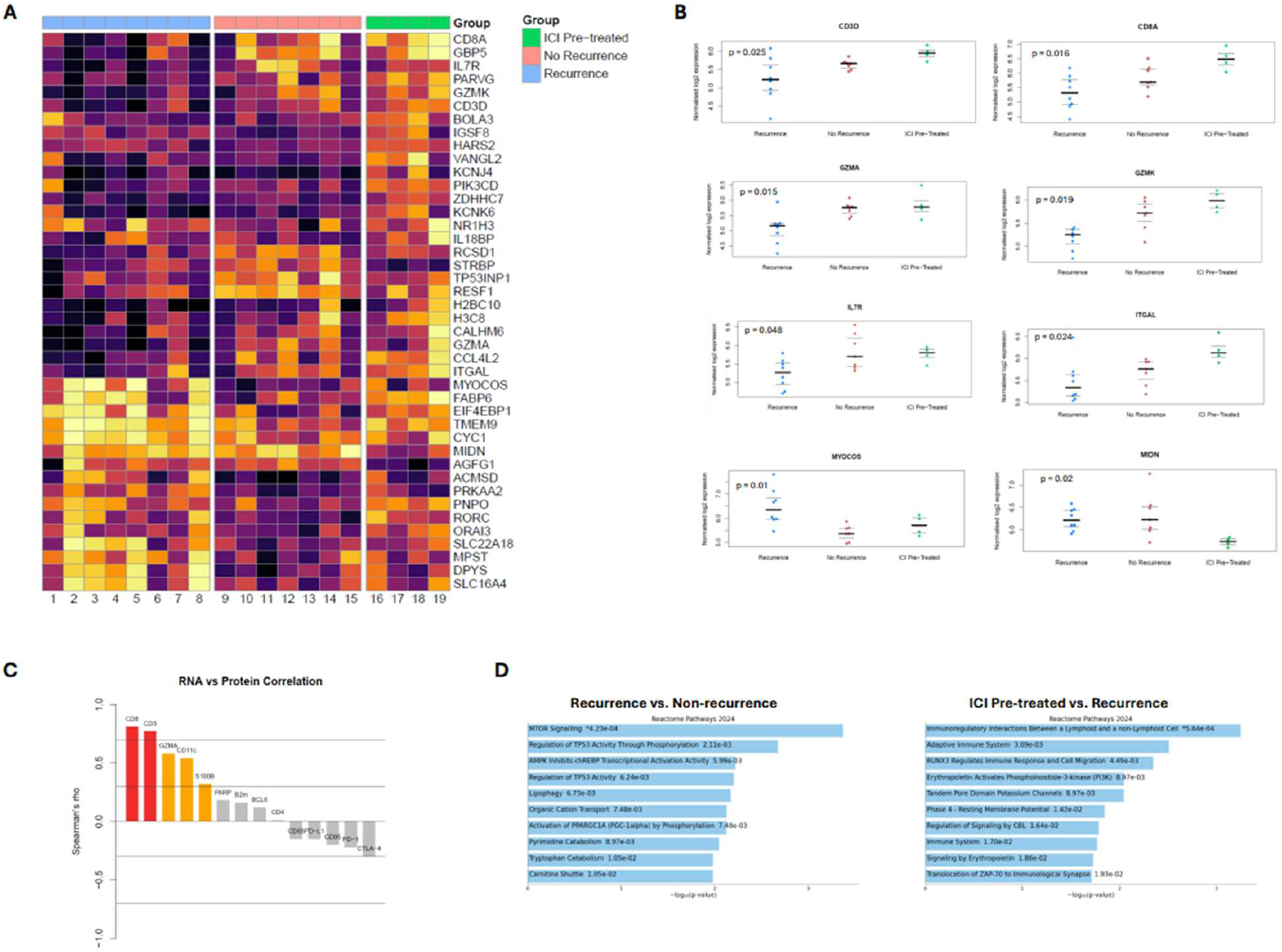
(A) Heatmap for RNA expression of genes that were most variably expressed between patients who were treatment-naïve with recurrence (blue), treatment-naïve without recurrence (pink), and with preoperative ICI exposure (green). (B) Normalized log2 RNA expression for selected genes that significantly differed across the three groups. (C) Plot of Spearman’s correlation coefficients for RNA and corresponding protein expression. (D) Reactome 2024 gene expression pathway comparing patients who were treatment-naïve with recurrence versus treatment-naïve without recurrence, as well as patients with preoperative ICI exposure versus treatment-naïve with recurrence.

Spearman’s correlations were determined between paired RNA and protein-product expression in matched tissues, and were found to be strongly correlated for CD8 (r=0.81, p<0.001) and CD3 (r=0.77, p<0.001), moderately correlated for GZMA (r=0.58, p=0.01), CD11c (r=0.54, p=0.02), and S100B (r=0.32, p=0.18), and poorly correlated for the remainder of the RNA-protein pairs (**Fig. 3C)**. Notably, clinically relevant immune checkpoints demonstrated weak negative correlations between their RNA gene expression and protein-product (PD-L1: r=-0.15, p=0.53; PD-1: r=-0.22, p=0.36; CTLA-4, r=-0.29, p=0.21).

Reactome 2024 gene expression pathway analysis was conducted using the between-groups differential expression analysis (**Fig. 3D)**. Tumor samples from treatment-naïve patients with recurrence had increased “MTOR Signaling” (p<0.001) and “Regulation of TP53 Activity Through Phosphorylation” (p=0.002) pathway enrichment, among others. Preoperative ICI tumor samples had increased enrichment of “Immunoregulatory Interactions Between a Lymphoid and a non-Lymphoid Cell” (p<0.001) and “Adaptive Immune System” (p=0.003) pathways, among others.

Gene expression data from the RNA analysis was utilized as input for immune cell deconvolution analysis using the *xCell* algorithm, as described by Aran and colleagues.^13^ Gene expression signatures were significantly different between groups for the immune cell subtypes CD8+ T-cell, CD8+ Tcm-cell, and CD8+ Tem-cell (**Fig. 4A)**. These three immune cell subtype signatures were significantly higher in the preoperative ICI and treatment-naïve without recurrence groups, and lower in treatment-naïve patients with recurrence (**Fig. 4B)**.

**Figure 4.**
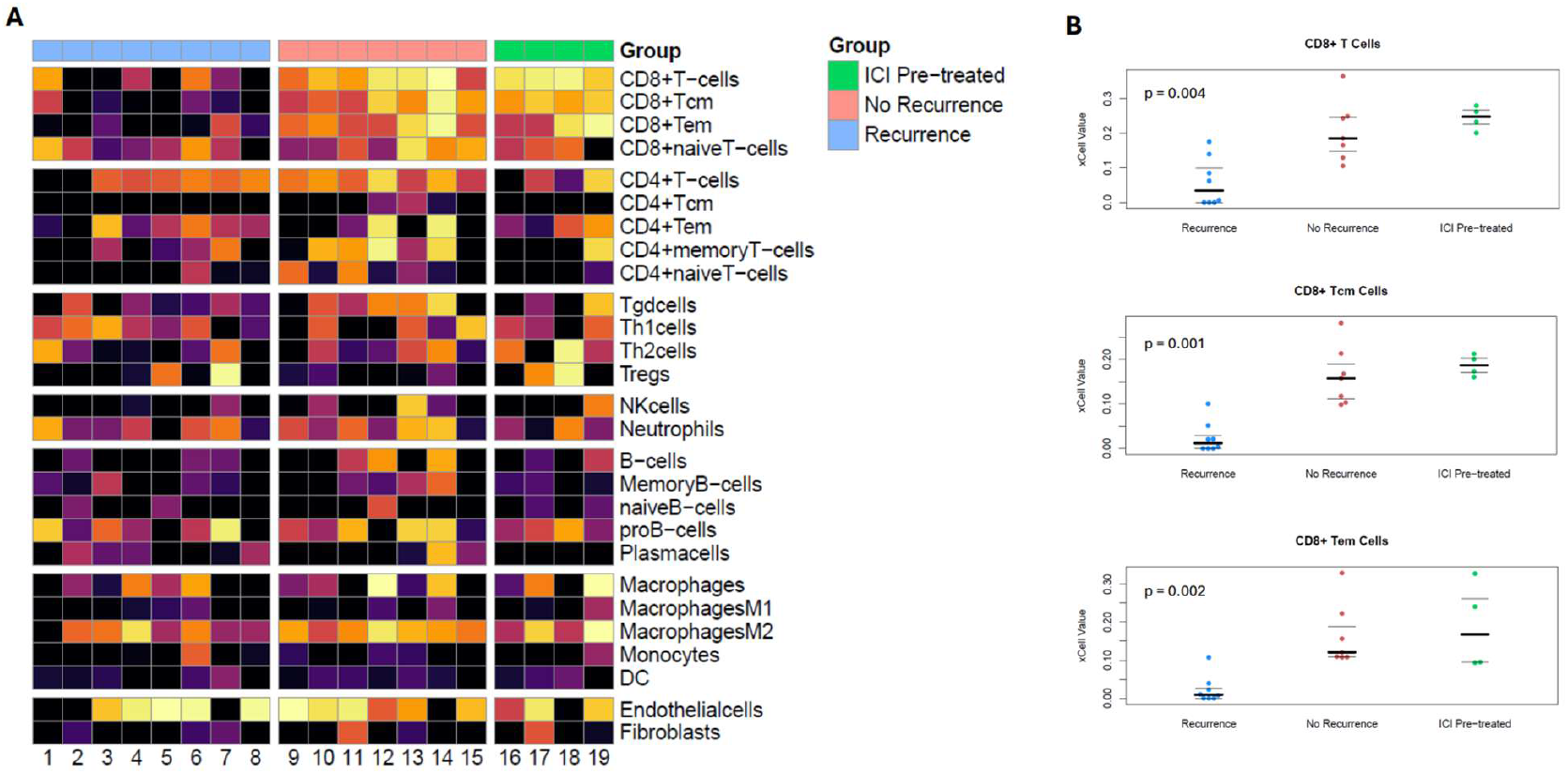
(A) Heatmaps for relative prevalence of immune cell subtypes based on *xCell* immune cell deconvolution applied to RNA expression data. (B) Jittered plots for relative prevalence of CD8+ T cells, CD8+ T_cm_ cells, and CD8+ T_em_ cells that show significant differences across the three groups (preoperative ICI, treatment-naive without recurrence, and treatment-naive with recurrence).

### Validation of gene expression results

We matched seven genes from our gene expression analysis associated with the preoperative ICI and treatment-naïve non-recurrence groups to RNA data from the NCT02210117 clinical trial (**Fig. 5A**). Further limiting this set of genes to only those that were significantly increased after ICI exposure yielded six genes (**Fig. 5B**). Finally, limiting this set to genes that had significantly increased expression in patients with objective response to therapy yielded four genes: *GZMK, GZMA, ITGAL*, and *IL7R*.

**Figure 5.**
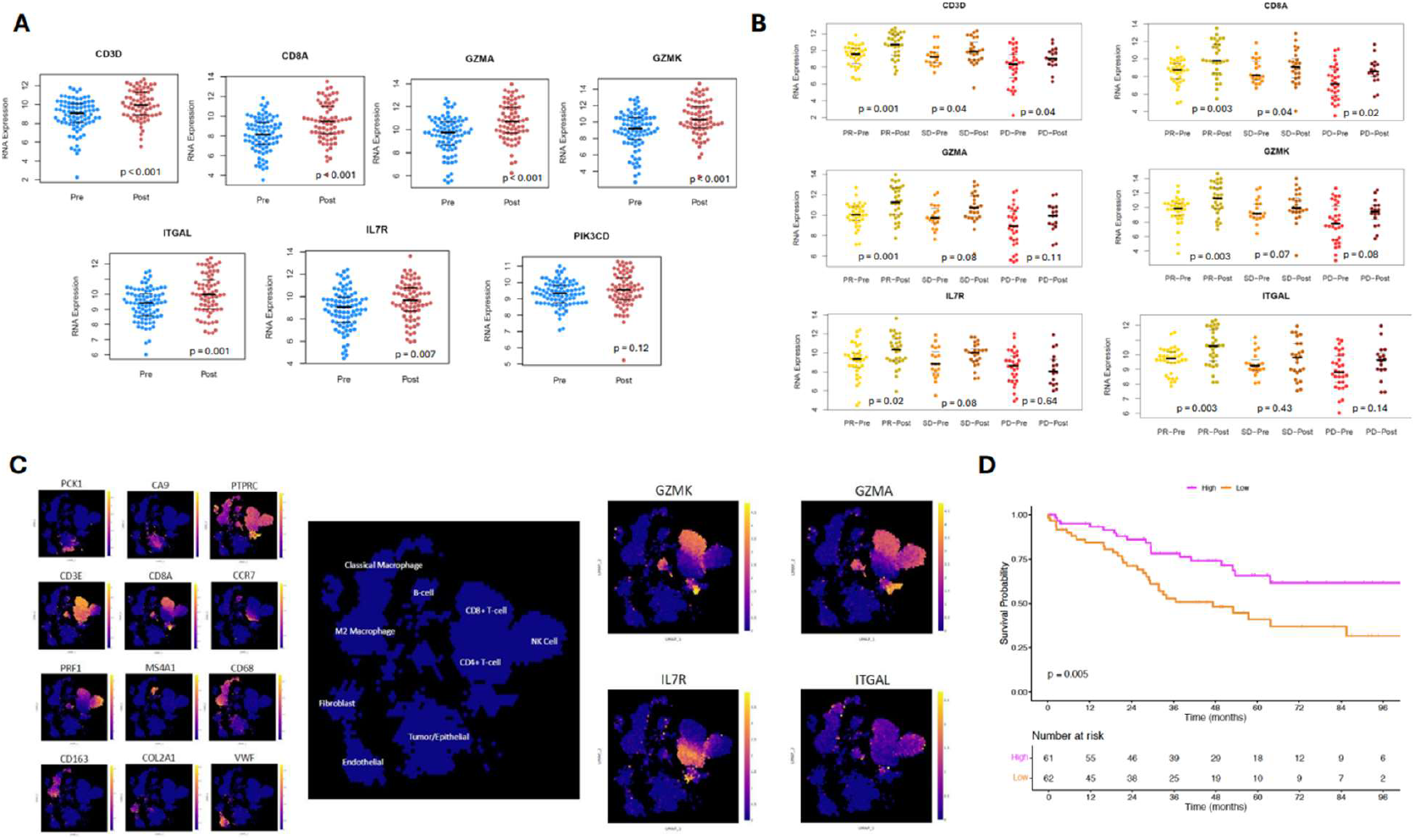
(A) Jittered plots of the 7 genes with significant differential expression in our study that were overlapped with RNA-seq data in the NCT02210117 clinical trial, based on pre- (blue) and post-ICI (red) exposure. (B) Jittered plots stratified by treatment response, for the 6 genes that showed significant pre-versus post-ICI differences. (C) Meta-analysis of single-cell RNA-seq data from ccRCC tumor tissue using the *UncoVer* platform; cell identities in the UMAP were determined using canonical gene markers and then expression of *GZMA, GZMK, IL7R*, and *ITGAL* was mapped to these clusters. (D). Kaplan-Meier curves for overall survival based on clinical stage III ccRCC patients from the TCGA KIRC cohort stratified by high (magenta) and low (orange) gene scores based on expression levels of *GZMA, GZMK, IL7R*, and *ITGAL*.

### External analysis of immune cell phenotype

On scRNA-seq meta-analysis of ccRCC tumors using *UncoVer*, there were distinct expression patterns based on different cell types noted (**Fig. 5C)**. We found that *GZMK* is primarily expressed in CD8+ T-cells, *GZMA* in CD8+ T-cells and NK cells, *IL7R* in CD4+ T-cells, and *ITGAL* in CD8+ T-cells, NK cells, and classical macrophages.

### Stratification of survival outcomes based on gene scores

The final gene set (*GZMK, GZMA, IL7R*, and *ITGAL*) was adapted into a gene score and applied to an independent cohort of treatment-naïve patients with stage-III ccRCC from the TCGA-KIRC cohort. Overall survival assessed using Kaplan-Meier estimates demonstrated significantly improved outcomes for patients with higher gene scores (log-rank p=0.005, **Fig. 5D)**.

## Discussion

We performed multimodal digital spatial analysis of stage III ccRCC primary tumors, finding that protein and RNA expression indicative of increased CD8+ Tcm and Tem cells was associated with a durable cancer-free state postoperatively. Interestingly, patients who received preoperative ICI treatment had similarly favorable findings, but without correspondingly improved clinical outcomes; three out of four of these patients experienced a recurrence during follow-up. We further investigated, validated, and refined our findings using several independent external cohorts, identifying *GZMK, GZMA, IL7R*, and *ITGAL* as genes that are modifiable with ICI therapy and associated with favorable clinical outcomes. As such, patients whose tumors have low baseline expression of these genes may benefit most from perioperative ICI therapy – a hypothesis that we intend to directly test in future experiments.

We reviewed the literature regarding these four genes with regards to their relation to RCC and malignancy more broadly. *GZMK* encodes the serine protease granzyme K, which is normally expressed by CD8+ T cells, natural killer cells, and innate-like T cells.^18^ Granzyme K facilitates apoptosis of target cells through single-stranded DNA nicks and damage to mitochondrial membranes.^19,20^ In the setting of RCC, clinical trials assessing PD-1 ICI in patients with advanced and metastatic RCC found that CD8+ T cells expressing *GZMK* were more prevalent in responders.^21,22^ Outside of RCC, TCGA analysis has also noted improved OS among patients with breast cancer and higher *GZMK* expression, and higher levels of *GZMK* have been noted in melanomas responsive to nivolumab compared to non-responders.^23^

*GZMA* encodes granzyme A and, like granzyme K, is a serine protease with tryptase activity that is prevalent in CD8+ T cells and natural killer cells.^24^ Within RCC, tumors with higher infiltration of natural killer cells had higher granzyme A expression, although this was not correlated with oncologic outcomes.^25^ *GZMA* expression was also associated with patients with localized ccRCC who had higher lymphocytic infiltration and T cell gene expression.^26^ Regarding prognosis, Matsushita and colleagues found that *GZMA* expression was not associated with OS among patients with ccRCC, though their cohort primarily consisted of Stage I and II tumors.^27^

*IL7R* encodes the alpha chain of IL-7R, a cell-surface receptor. This receptor plays an important role in lymphocyte development and can promote cell survival through activation of the JAK/STAT signaling pathway.^28^ In a mouse model with a chronic viral infection, PD-1 blockade led to increased *IL7R* expression and IL-7 signaling, which in term was associated with transient reinvigoration of exhausted CD8+ T cells.^29^ The literature for this gene’s relation to RCC is relatively sparse, with increased proliferation of tumor-infiltrating lymphocytes observed with IL-7 treatment.^30^

*ITGAL* codes for integrin alpha L chain, which is a subunit of the lymphocyte function-associated antigen-1 (LFA-1) and is critical for leukocyte migration. RCC cancers overexpress *ITGAL* compared to normal control tissue and greater expression is associated with higher tumor grade. That same study also found that as part of a 10-gene expression signature, greater *ITGAL* expression predicted poorer survival using TCGA data.^31^ The study did not stratify *ITGAL* expression within stages in relation to survival though. Outside of RCC, lower ITGAL expression in non-small-cell lung cancer was associated with worse prognosis, with higher immune infiltration in malignant tissues.^32^ Similar trends were seen in patients with melanoma.^33^

Within the protein expression analysis, the preoperative ICI group had the highest levels of CD11c, an integrin protein primarily expressed on dendritic cells. Prior analysis of gene expression profiles from the Gene Expression Omnibus (GEO) database found that *ITGAX*, which codes for CD11c, is upregulated in ccRCC tissue and associated with poorer prognosis.^34^ In vitro studies of ovarian tumor cells have suggested that *ITGAX* overexpression may activate the PI3k/Akt pathway.^35,36^ Recent spatial transcriptomic studies have consistently identified integrins as being biologically meaningful components of the tumor immune microenvironment.^37^

The assumption is often made that expression of a gene and its downstream protein product will be highly correlated, but this was not the case in our data. Of the 14 matched gene-protein pairings, only two had a strong positive correlation (CD8 and CD3), and three had a moderately positive correlation (GZMA, CD11c, and S100B). It is particularly noteworthy that the immune checkpoint proteins PD-L1, PD-1, and CTLA-4 all demonstrated weak negative correlations, which has previously been noted in the literature.^38–41^ This data underscores the point that protein and RNA data are not interchangeable, especially for immune checkpoints.

### Limitations

There were several limitations to this study. Primarily, our cohort was small (n=19 patients), though we validated these findings using larger independent studies. This experiment was designed to favor a deep and comprehensive molecular analysis of fewer specimens, as opposed to a superficial analysis of many specimens. Future studies validating these findings will be more targeted and feature a larger sample size. Additionally, lack of single-cell resolution limits immune cell phenotyping to relatively broad subtypes, limiting analysis of granular subtypes of exhausted T-cells and anti-inflammatory macrophages that have previously been described as important in ccRCC translational studies.^42–44^ Additionally, we did not have protein or RNA expression data that was spatially resolved at the single-cell level, so could not determine CD8+ clustering patterns that better describe immune infiltration.^39^ Ultimately, the described findings absolutely require further study and validation before being directly applied to clinical practice.

## Conclusions

Multimodal digital spatial molecular analysis identified that patients with Stage III ccRCC who received pre-operative ICI had increased intratumoral expression of genes and protein products indicative of CD8+ T-cell effector and central-memory phenotypes – similar to untreated patients who did not recur after surgery. Despite this favorable immune composition, our ICI-pretreated patients recurred at similar rates to untreated patients. Overall, receipt of preoperative ICI appeared to favorably alter the tumor immune microenvironment but did not result in better outcomes.

Further refinement of our analysis using NCT02210117 clinical trial data suggested that increased gene expression of *GZMK, GZMA, ITGAL*, and *IL7R* is inducible with ICI therapy and associated with favorable clinical outcomes. Patients whose tumors have low initial expression of these genes may derive the most benefit from perioperative ICI. Once published, gene expression data from the PROSPER and KEYNOTE-564 trials will be critical to test this hypothesis.

## Supporting information

Supplemental Table 1 (updated)

## Data Availability

All data produced in the present study are available upon reasonable request to the authors.

## Acknowledgements

We thank the patients and families who participated in this research, the OHSU Knight Cancer Center Bio-library for specimen management, and the Nanostring Technologies team for technical support with digital spatial profiling.

## References

1. Levy DA, Slaton JW, Swanson DA, Dinney CPN. STAGE SPECIFIC GUIDELINES FOR SURVEILLANCE AFTER RADICAL NEPHRECTOMY FOR LOCAL RENAL CELL CARCINOMA. The Journal of Urology. 1998;159(4):1163–1167. doi:10.1016/S0022-5347(01)63541-9

2. Monteiro FSM, Soares A, Rizzo A, et al. The role of immune checkpoint inhibitors (ICI) as adjuvant treatment in renal cell carcinoma (RCC): A systematic review and meta-analysis. Clinical Genitourinary Cancer. 2023;21(3):324–333. doi:10.1016/j.clgc.2023.01.005

3. Choueiri TK, Tomczak P, Park SH, et al. Overall Survival with Adjuvant Pembrolizumab in Renal-Cell Carcinoma. New England Journal of Medicine. 2024;390(15):1359–1371. doi:10.1056/NEJMoa2312695

4. kidney.pdf. Accessed May 1, 2025. https://www.nccn.org/professionals/physician_gls/pdf/kidney.pdf

5. Allaf ME, Kim SE, Master V, et al. Perioperative nivolumab versus observation in patients with renal cell carcinoma undergoing nephrectomy (PROSPER ECOG-ACRIN EA8143): an open-label, randomised, phase 3 study. Lancet Oncol. 2024;25(8):1038–1052. doi:10.1016/S1470-2045(24)00211-0

6. Fropf R, Griswold M, Zimmerman S, et al. The GeoMx® Human Whole Transcriptome Atlas for the Digital Spatial Profiler:

7. Hernandez S, Lazcano R, Serrano A, et al. Challenges and Opportunities for Immunoprofiling Using a Spatial High-Plex Technology: The NanoString GeoMx® Digital Spatial Profiler. Front Oncol. 2022;12:890410. doi:10.3389/fonc.2022.890410

8. Whitepaper Archives. NanoString. 2025. Accessed June 10, 2025. https://nanostring.com/em_resources_type/whitepaper/

9. FL_MK2295_nCounter_Grant_Support_R5.pdf. Accessed May 2, 2025. https://nanostring.com/wp-content/uploads/FL_MK2295_nCounter_Grant_Support_R5.pdf

10. Law CW, Chen Y, Shi W, Smyth GK. voom: precision weights unlock linear model analysis tools for RNA-seq read counts. Genome Biology. 2014;15(2):R29. doi:10.1186/gb-2014-15-2-r29

11. Ritchie ME, Phipson B, Wu D, et al. limma powers differential expression analyses for RNA-sequencing and microarray studies. Nucleic Acids Research. 2015;43(7):e47. doi:10.1093/nar/gkv007

12. Milacic M, Beavers D, Conley P, et al. The Reactome Pathway Knowledgebase 2024. Nucleic Acids Research. 2024;52(D1):D672–D678. doi:10.1093/nar/gkad1025

13. Aran D, Hu Z, Butte AJ. xCell: digitally portraying the tissue cellular heterogeneity landscape. Genome Biol. 2017;18(1):220. doi:10.1186/s13059-017-1349-1

14. Kolde R. raivokolde/pheatmap. Published online June 10, 2025. Accessed June 10, 2025. https://github.com/raivokolde/pheatmap

15. Goswami S, Gao J, Basu S, et al. Immune checkpoint inhibitors plus debulking surgery for patients with metastatic renal cell carcinoma: clinical outcomes and immunological correlates of a prospective pilot trial. Nat Commun. 2025;16(1):1846. doi:10.1038/s41467-025-57009-z

16. Eisenhauer EA, Therasse P, Bogaerts J, et al. New response evaluation criteria in solid tumours: revised RECIST guideline (version 1.1). Eur J Cancer. 2009;45(2):228–247. doi:10.1016/j.ejca.2008.10.026

17. Lecuyer GCV, Lardenois A, Chalmel F. UncoVer: A Web-based Resource for Single-cell and Spatially Resolved Omics Data in Uro-oncology. European Urology Oncology. 2024;7(6):1545–1547. doi:10.1016/j.euo.2024.04.008

18. Donado CA, Theisen E, Zhang F, et al. Granzyme K activates the entire complement cascade. Nature. 2025;641(8061):211–221. doi:10.1038/s41586-025-08713-9

19. Zhao T, Zhang H, Guo Y, et al. Granzyme K cleaves the nucleosome assembly protein SET to induce single-stranded DNA nicks of target cells. Cell Death Differ. 2007;14(3):489–499. doi:10.1038/sj.cdd.4402040

20. Zhao T, Zhang H, Guo Y, Fan Z. Granzyme K Directly Processes Bid to Release Cytochrome c and Endonuclease G Leading to Mitochondria-dependent Cell Death*. Journal of Biological Chemistry. 2007;282(16):12104–12111. doi:10.1074/jbc.M611006200

21. Au L, Hatipoglu E, Robert de Massy M, et al. Determinants of anti-PD-1 response and resistance in clear cell renal cell carcinoma. Cancer Cell. 2021;39(11):1497-1518.e11. doi:10.1016/j.ccell.2021.10.001

22. Gu L, Peng C, Liang Q, et al. Neoadjuvant toripalimab plus axitinib for clear cell renal cell carcinoma with inferior vena cava tumor thrombus: NEOTAX, a phase 2 study. Signal Transduct Target Ther. 2024;9:264. doi:10.1038/s41392-024-01990-2

23. Li Z, Xie Q, Zhao F, et al. Exploring GZMK as a prognostic marker and predictor of immunotherapy response in breast cancer: unveiling novel insights into treatment outcomes. J Cancer Res Clin Oncol. 2024;150(6):1–21. doi:10.1007/s00432-024-05791-6

24. Martinvalet D, Zhu P, Lieberman J. Granzyme A Induces Caspase-Independent Mitochondrial Damage, a Required First Step for Apoptosis. Immunity. 2005;22(3):355–370. doi:10.1016/j.immuni.2005.02.004

25. Schleypen JS, Baur N, Kammerer R, et al. Cytotoxic Markers and Frequency Predict Functional Capacity of Natural Killer Cells Infiltrating Renal Cell Carcinoma. Clinical Cancer Research. 2006;12(3):718–725. doi:10.1158/1078-0432.CCR-05-0857

26. Ghatalia P, Gordetsky J, Kuo F, et al. Prognostic impact of immune gene expression signature and tumor infiltrating immune cells in localized clear cell renal cell carcinoma. J Immunother Cancer. 2019;7(1):139. doi:10.1186/s40425-019-0621-1

27. Matsushita H, Sato Y, Karasaki T, et al. Neoantigen Load, Antigen Presentation Machinery, and Immune Signatures Determine Prognosis in Clear Cell Renal Cell Carcinoma. Cancer Immunol Res. 2016;4(5):463–471. doi:10.1158/2326-6066.CIR-15-0225

28. Mazzucchelli R, Durum SK. Interleukin-7 receptor expression: intelligent design. Nat Rev Immunol. 2007;7(2):144–154. doi:10.1038/nri2023

29. Pauken KE, Sammons MA, Odorizzi PM, et al. Epigenetic stability of exhausted T cells limits durability of reinvigoration by PD-1 blockade. Science. 2016;354(6316):1160–1165. doi:10.1126/science.aaf2807

30. Ditonno P, Tso CL, Sakata T, deKernion JB, Belldegrun A. Regulatory effects of interleukin-7 on renal tumor infiltrating lymphocytes. Urol Res. 1992;20(3):205–210. doi:10.1007/BF00299718

31. Boguslawska J, Kedzierska H, Poplawski P, Rybicka B, Tanski Z, Piekielko-Witkowska A. Expression of Genes Involved in Cellular Adhesion and Extracellular Matrix Remodeling Correlates with Poor Survival of Patients with Renal Cancer. The Journal of Urology. Published online June 2016. doi:10.1016/j.juro.2015.11.050

32. Zhang R, Zhu G, Li Z, et al. ITGAL expression in non-small-cell lung cancer tissue and its association with immune infiltrates. Front Immunol. 2024;15:1382231. doi:10.3389/fimmu.2024.1382231

33. Deng T, Wang C, Gao C, Zhang Q, Guo J. ITGAL as a prognostic biomarker correlated with immune infiltrates in melanoma. Front Oncol. 2023;13:1181537. doi:10.3389/fonc.2023.1181537

34. Sui Y, Lu K, Fu L. Prediction and analysis of novel key genes ITGAX, LAPTM5, SERPINE1 in clear cell renal cell carcinoma through bioinformatics analysis. PeerJ. 2021;9:e11272. doi:10.7717/peerj.11272

35. Wang J, Yang L, Liang F, Chen Y, Yang G. Integrin alpha x stimulates cancer angiogenesis through PI3K/Akt signaling–mediated VEGFR2/VEGF-A overexpression in blood vessel endothelial cells. Journal of Cellular Biochemistry. 2019;120(2):1807–1818. doi:10.1002/jcb.27480

36. Williams KA, Lee M, Hu Y, et al. A Systems Genetics Approach Identifies CXCL14, ITGAX, and LPCAT2 as Novel Aggressive Prostate Cancer Susceptibility Genes. PLOS Genetics. 2014;10(11):e1004809. doi:10.1371/journal.pgen.1004809

37. Soupir AC, Hayes MT, Peak TC, et al. Increased spatial coupling of integrin and collagen IV in the immunoresistant clear-cell renal-cell carcinoma tumor microenvironment. Genome Biol. 2024;25:308. doi:10.1186/s13059-024-03435-z

38. Chakiryan NH, Hajiran A, Kim Y, et al. Correlating Immune Cell Infiltration Patterns with Recurrent Somatic Mutations in Advanced Clear Cell Renal Cell Carcinoma. Eur Urol Focus. 2022;8(3):784–793. doi:10.1016/j.euf.2021.04.014

39. Chakiryan NH, Kim Y, Berglund A, et al. Geospatial characterization of immune cell distributions and dynamics across the microenvironment in clear cell renal cell carcinoma. J Immunother Cancer. 2023;11(4):e006195. doi:10.1136/jitc-2022-006195

40. Zerdes I, Karafousia V, Mezheyeuski A, et al. Discordance of PD-L1 Expression at the Protein and RNA Levels in Early Breast Cancer. Cancers. 2021;13(18):4655. doi:10.3390/cancers13184655

41. Nicolet BP, Wolkers MC. The relationship of mRNA with protein expression in CD8+ T cells associates with gene class and gene characteristics. PLOS ONE. 2022;17(10):e0276294. doi:10.1371/journal.pone.0276294

42. Braun DA, Street K, Burke KP, et al. Progressive immune dysfunction with advancing disease stage in renal cell carcinoma. Cancer Cell. 2021;39(5):632-648.e8. doi:10.1016/j.ccell.2021.02.013

43. Bi K, He MX, Bakouny Z, et al. Tumor and immune reprogramming during immunotherapy in advanced renal cell carcinoma. Cancer Cell. 2021;39(5):649-661.e5. doi:10.1016/j.ccell.2021.02.015

44. Krishna C, DiNatale RG, Kuo F, et al. Single-cell sequencing links multiregional immune landscapes and tissue-resident T cells in ccRCC to tumor topology and therapy efficacy. Cancer Cell. 2021;39(5):662-677.e6. doi:10.1016/j.ccell.2021.03.007

